# Multi-voxel neuro-reinforcement changes resting-state functional connectivity: A pilot study

**DOI:** 10.1101/2023.11.10.23298400

**Authors:** Shawn Wang, Cody A. Cushing, Hakwan Lau, Michelle G. Craske, Vincent Taschereau-Dumouchel

## Abstract

**Background:** Multi-voxel neuro-reinforcement has been shown to selectively reduce amygdala reactivity in response to feared stimuli, but the precise mechanisms supporting these effects are still unknown. The current pilot study seeks to identify potential intermediaries of change using functional brain connectivity at rest.

**Methods:** Individuals (N = 11) diagnosed with at least two animal subtype specific phobias took part in a double-blind multi-voxel neuro-reinforcement clinical trial targeting one of two phobic animals, with the untargeted animal as placebo control. Changes in whole-brain resting state functional connectivity from pre-treatment to post-treatment were measured using group ICA. These changes were tested to see if they predicted the previously observed decreases in amygdala reactivity in response to images of target phobic animals.

**Results:** A common functional connectivity network overlapping with the visual network was identified in resting state data pre-treatment and post-treatment. Significant increases in functional connectivity in this network from pre-treatment to post-treatment were found in higher level visual and cognitive processing regions of the brain. Increases in functional connectivity in these regions also significantly predicted decreases in task-based amygdala reactivity to targeted phobic animals following multi-voxel neuro-reinforcement. Specifically, greater increases of functional connectivity pre-treatment to post-treatment were associated with greater decreases of amygdala reactivity to target phobic stimuli pre-treatment to post-treatment.

**Conclusions:** These findings provide preliminary evidence that multi-voxel neuro-reinforcement can induce persisting functional connectivity changes in the brain. Moreover, these changes in functional connectivity were not limited to the direct area of neuro-reinforcement, suggesting neuro-reinforcement may change how the targeted region interacts with other brain regions. Identification of these brain regions represent a first step towards explaining the underlying mechanisms of change in previous multi-voxel neuro-reinforcement studies. Future research should seek to replicate these effects in a larger sample size to further assess their role in the effects observed from multi-voxel neuro-reinforcement.

---

Exposure therapy is one of the most effective and empirically validated forms of treatment for anxiety disorders (Norton & Price, 2007), including specific phobia (Hirai et al., 2007). However, because exposure relies on confronting aversive stimuli and can induce intense fear, many of those suffering from anxiety disorders terminate treatment early (Loerinc et al., 2015) or do not even begin treatment (Wang et al., 2000, 2005), leaving a large proportion of this population without adequate help.

To circumvent the aversive nature of traditional exposure therapy, multi-voxel neuro-reinforcement has been explored as an alternative intervention (Cushing, Lau, et al., 2023; Koizumi et al., 2016; Taschereau-Dumouchel et al., 2018). This method uses closed-loop real-time functional magnetic resonance imaging (fMRI) to pair monetary reward with nonconscious neural activation of a multi-voxel category representation (e.g. spider) (Koizumi et al., 2016; Shibata et al., 2011; Shibata et al., 2019; Watanabe et al., 2017). In doing so, participants remain unaware of the nature of the reinforced representation and thus avoid the discomfort normally associated with exposure therapy. Previous experiments with multi-voxel neuro-reinforcement for feared stimuli have demonstrated its ability to decrease amygdala reactivity to feared animals whose category patterns were targeted with multi-voxel neuro-reinforcement (Taschereau-Dumouchel et al., 2018).

Recently, we conducted a randomized, double-blind clinical trial of a diagnosed sample with animal phobia (Cushing, Lau, et al., 2023), focusing on amygdala reactivity in response to images of feared stimuli as the primary outcome (Koizumi et al., 2016; Taschereau-Dumouchel et al., 2018). Participants with at least two diagnosed animal phobias underwent multi-voxel neuro-reinforcement with one animal phobia as a target for treatment and another one as a double-blind placebo control condition. During neuro-reinforcement, participants received monetary reward as a function of the activation of the target animal category pattern in the ventral temporal cortex (VT). They could track their performance through visual feedback in the form of a growing or shrinking disc. Activation of category representations was determined using a machine-learning classifier trained to predict the presence of the target category from multi-voxel brain activity in VT. This brain region was chosen as the location for reinforcement for its role in high-level categorization of visual stimuli (Grill-Spector & Weiner, 2014) and use in previous experiments (Taschereau-Dumouchel et al., 2018). Importantly, the activation of this representation was implicit and nonconscious as participants were not given specific instruction on what the feedback represented in their brain. Participants were unaware of the purpose of the neuro-reinforcement other than it being a treatment and participant strategies were checked daily to ensure participants did not attach the visual feedback to the targeted animal category.

Our trial found significantly greater reductions in amygdala reactivity for the targeted phobia compared to the placebo control (Cushing, Lau, et al., 2023). This result lines up with previous work (Koizumi et al., 2016; Taschereau-Dumouchel et al., 2018) that demonstrate similar reductions in amygdala responses. However, the mechanisms of these effects from neuro-reinforcement remain unclear (Shibata et al., 2019; Taschereau-Dumouchel, Kawato, et al., 2020). Some nascent research has focused on functional connectivity during resting state as a means to uncover intermediary changes from neuro-feedback (Megumi et al., 2015; Scheinost et al., 2013). One preliminary investigation conducted a seed-based analysis of relationships between resting state functional connectivity and the prefrontal cortex following a multi-voxel neuro-reinforcement intervention (Taschereau-Dumouchel, Chiba, et al., 2020). Here, we are building upon these findings in a pilot study by conducting an investigation in a population with diagnosed animal phobia. More specifically, we examined resting state functional connectivity changes as a result of multi-voxel neuro-reinforcement and investigated their association with previously observed effects. Using data collected as part of Cushing et al., (2023), we conducted a Group-ICA functional connectivity analysis of resting state fMRI data (Calhoun et al., 2009) to characterize whole-brain functional connectivity networks in a data-driven fashion. We then examined how these networks changed following neuro-reinforcement and related them to previously observed effects in the amygdala (Cushing, Lau, et al., 2023).

Previous research examining information transmission during multi-voxel neuro-reinforcement has demonstrated that information is transmitted between the targeted region VT and the lingual gyrus, inferior temporal gyrus, and fusiform gyrus during reinforcement trials (Taschereau-Dumouchel et al., 2018). Importantly, this effect was not found for the amygdala where effects are found following neuro-reinforcement. These findings suggest that neural activation during multi-voxel neuro-reinforcement is relatively localized within regions of the brain responsible for higher level visual processing, rather than acting on affective regions like the amygdala directly. Consequently, while this transmission was observed during multi-voxel neuro-reinforcement, we predict we will observe longer-term functional connectivity changes outside the neuro-reinforcement task at rest in these areas due to their engagement during the task. Therefore, we hypothesize changes in functional connectivity during rest in areas of the brain involved in high level visual processing and not in the amygdala as a result of repeated multi-voxel pattern reinforcement in the VT. Secondly, following our main findings (Cushing, Lau, et al., 2023), we hypothesize changes in resting state connectivity to be associated with decreased amygdala reactivity to the target phobia observed in the fear task. This association would support any observed functional connectivity changes as potentially contributing to the mechanisms of previously observed neuro-reinforcement effects.

## METHODS

### Participant Screening

Participants were recruited through flyers, listings on UCLA campus newsletters, and advertisements on online boards (e.g. Nextdoor, Reddit, etc). Participants were screened using a modified Fear Survey Schedule (Wolpe & Lang, 1964) and were required to endorse fear levels of at least 3 (0-8 point scale) for at least two animals available in our image dataset to be eligible. Participants meeting these criteria were then interviewed using the ADIS-V (Brown & Barlow, 2014) by a trained staff member and reviewed for consensus with the Principal Investigator (Craske). During the ADIS-V interview, diagnoses were ascertained through interviewer ratings of fear and avoidance using 0 (no fear/never avoids) to 8 (very severe fear/always avoids) point scales; participants were required to receive ratings of at least 4 for either fear or avoidance for at least two specific animal phobias. Interviewers also provided ratings of clinical severity on a 0 - 8 point scale, taking into account symptom severity, distress, and impairment of each phobic animal (0 = no symptoms, distress, and impairment, 8 = very severe symptoms, distress, and impairment). Each phobia was required to have at least a mild clinical severity rating (2 or above). Participants were excluded if at any time they 1) were unable to understand or complete the informed consent process; 2) were unable to respond to screening questions; 3) did not have normal/corrected to normal vision/hearing; 4) had history of neurological disease/defect or any other serious and unstable medical conditions; 5) were diagnosed with current MDD, SUD, OCD, PTSD, bipolar disorder, or psychosis; 6) were currently taking psychotropic medication, or; 7) did not meet MRI scanning safety criteria.

After informed consent as approved by the Institutional Review Board of the University of California, Los Angeles, 23 participants (mean age (s.d.) = 26.27 (10.87), 81.82% female) who were diagnosed through clinical interviewing for at least two animal phobias were enrolled to undergo multi-voxel neuro-reinforcement. Participants were randomly assigned to 1, 3, or 5 days of multi-voxel neuro-reinforcement. From the 23 participants, two were unable to complete the full treatment (1 from scheduling issues, the other from technical issues). Out of the remaining 21 who completed treatment, one was excluded because they reported nausea during the procedure and two did not complete the “fear test” amygdala response task (1 could not keep eyes open during task due to fear, 1 from technical issues), leaving 18 participants with complete amygdala response data who were included in the report by Cushing et al. (2023). However, 7 of these 18 participants did not have complete pre-post resting state scan data (6 due to scanning time limitations, 1 fell asleep during scan), leaving a total of 11 participants analyzed. The 11 participants reflect not only the aforementioned technical and logistical difficulties but also the immense challenges in recruitment presented by the COVID-19 pandemic which occurred in the middle of the study and severely hampered both in-person study data collection and interest in research participation.

### Multi-voxel Neuro-reinforcement

Participants were randomized to either 1, 3, or 5 sessions of neuro-reinforcement prior to beginning pre-treatment. For each participant, one of their diagnosed phobias was chosen to be the “target” animal to be reinforced, and another was chosen to be the “control” animal, which acted as a placebo and was not reinforced (Cushing, Lau, et al., 2023). Neither target nor control animal were known by the participant nor the experimenter, allowing us to maintain a double-blind. During neuro-reinforcement sessions, participants were asked to “[think about] whatever [they] can to try and get the best score”, during which a neuro-reinforcement method (Cushing, Lau, et al., 2023; Taschereau-Dumouchel et al., 2018) rewarded them based on successful activation of their phobic image category.

Participants’ scores were represented to them as a green disk, with larger disks indicating higher scores. The score participants received directly corresponded to the real-time output of the sparse multinomial logistic regression decoder constructed for that participant’s target phobia (Cushing, Lau, et al., 2023). They were also told that the size of the disk corresponded to the amount of their monetary reward at the end of the run, with their reward based on the average feedback score during the run (e.g. 50% average score meant 50%, or $3.00, of the $6.00 maximum payout per run) Additionally, if participants were able to achieve above an 80% score for three trials in a row, they were told they would receive a high score streak bonus of $2.00.

### Pre/Post Neuro-reinforcement Assessments

Phobic participants completed pre- and post-treatment fMRI assessments in which they completed a “fear test” task while their BOLD signals were recorded. The fear test presented images (6s) to participants belonging to either their target phobic animal, control phobic animal, or a randomly selected non-phobic animal or object as determined by diagnostic interview. They then were prompted to rate how fearful they found the image on a 7-point Likert scale. Full details of the fear task are reported in (Cushing, Lau, et al., 2023).

### fMRI Data Collection

MRI scans were acquired on a 3T Siemens Prisma scanner with a 32-channel head coil at the UCLA Ahmanson-Lovelace Brain Mapping Center.

Resting state scans were collected both at participants’ initial decoder construction visit before beginning pre-treatment and at their final post-treatment assessment visit. Data were collected using a multi-band sequence in the posterior (P) to anterior (A) direction with an acceleration factor of 8. Voxel size was 2.0 x 2.0 x 2.0 mm ^3^ with a field of view of 208 x 208 mm ^2^. Images were collected from 72 interleaved slices with a TR of 800 ms, TE of 37.00 ms, and a flip angle of 52 degrees. Opposite direction phase-encoded spin echo field maps were collected before functional runs to be used for distortion correction.

Fear test scans for the amygdala analysis were collected at pre-treatment and post-treatment visits. Data were collected using a multi-band sequence from posterior (P) to anterior (A) and with an acceleration factor of 8. The voxel size was 2.0 x 2.0 x 2.0 mm^3^ with a field of view of 208×208mm^2^. Images were collected from 72 interleaved slices with a TR of 800ms, TE of 37.00 ms, and flip angle of 52 degrees. During the COVID-19 shutdown, this specific sequence was modified to better capture BOLD activity in subcortical structures like the amygdala. Details of this sequence can be found in Cushing et al., (2023). The new scanning sequence had no effect on overall Beta estimates for the amygdala in the fear test (*t*(16)=-0.391, p=0.70), suggesting it is unlikely that any findings in the study were a result of this sequence change during COVID. Opposite direction phase-encoded spin echo field maps were also collected before functional scans in order to be used for distortion correction.

Anatomical scans came from a T1-weighted sequence with volumetric navigators (vNAV) with prospective motion correction (TR: 2500ms/TI: 1000ms/Flip Angle: 8.0 degrees/Voxel Size: 0.8×0.8×0.8mm/Matrix Size: 256×256/Num. Slices: 208/Slice Thickness: 0.8mm).

### Amygdala Processing

Fear test fMRI data were distortion corrected using opposite phase-encoded direction spin echo field map sequences with FSL topup (Andersson et al., 2003; Smith et al., 2004). Next, brain extraction of the T1 anatomical image was completed using bet (Smith, 2002). Preprocessing and ICA-decomposition were completed using FSL melodic and FEAT (*FMRIB’s Software Library*, n.d.), where data were motion corrected through mcflirt (Jenkinson et al., 2002), brain extracted using bet (Smith, 2002), spatially smoothed with a Gaussian kernel of FWHM 4.0mm, then intensity normalized and highpass filtered with a gaussian-weighted least squares straight line fitting with sigma=50.0s. Registration to the standard MNI space was performed using FLIRT and refined through nonlinear registration using FNIRT (Jenkinson et al., 2002; Jenkinson & Smith, 2001). A high-contract single-band reference image collected at the beginning of each run allowed for an improved registration of the multi-band images.

ICA components were visually inspected and manually cleaned to remove movement artifacts and other noise. Artifact Detection Tools (ART, https://www.nitrc.org/projects/artifact_detect) was used to further account for movement by generating motion regressors and identify outlier timepoints to be censored. Next, first-level GLMs were calculated using SPM12 with a temporal derivative to control for slice-timing differences. The target phobia, control phobia, neutral animal, and neutral object image presentation onsets were fit as regressors with a duration of 0 seconds to model event-related responses. Similar to previous methods (Cushing, Lau, et al., 2023; Koizumi et al., 2016; Taschereau-Dumouchel et al., 2018), BOLD data from only the first two trials of each target phobia and control phobia image were analyzed.

Freesurfer segmentation of the T1 anatomical image created bilateral amygdala masks which were then transformed into the participant’s native functional space. We used marsbar (Brett et al., 2002) to extract average parameter estimates from the amygdala masks. The phobic animal estimates were then baseline corrected by subtracting the average amygdala response to the neutral animal from the target and control phobias within runs. Finally, these corrected amygdala responses were averaged across runs for both pre/post assessments.

### Preprocessing

Resting state scans were first distortion corrected using FSL topup (Andersson et al., 2003; Smith et al., 2004) with opposite direction phase-encoded spin echo field maps. Brain extraction of the anatomical image was conducted using bet (Smith, 2002). Preprocessing was conducted using FSL FEAT (FMRIB’s Software Library, www.fmrib.ox.ac.uk/fsl). Using FEAT, data were motion corrected using mcflirt (Jenkinson et al., 2002) and brain extracted with bet (Smith, 2002). We spatial smoothed with a Gaussian kernel of FWHM of 4.0. No highpass/lowpass filtering occurred at this stage in anticipation of using ICA-AROMA. A bandpass filter was applied at a later stage. The functional scans were then registered to the standard MNI space and refined with nonlinear registration using FLIRT and FNIRT (Jenkinson et al., 2002; Jenkinson & Smith, 2001), respectively. A high-contrast single-band reference image collected at the beginning of each resting state scan enabled better registration with standard space templates.

Next, motion in the data was accounted for using ICA-AROMA (FSL Automatic Removal Of Motion Artifacts) (Pruim et al., 2015) and anatomical segmentation was conducted using FSL FAST (Zhang et al., 2001). Afterwards, additional motion and physiological (i.e. white matter, CSF) noise were regressed out of the data using the 3dTproject function from the AFNI toolbox (Cox, 1996; Cox & Hyde, 1997). A bandpass filter was also applied to remove frequencies below .008 and above .08 and then the filtered data were transformed into the standard space using the FSL applywarp function (Jenkinson et al., 2012).

### Independent Component Analysis (ICA)

Group-ICA was conducted using FSL MELODIC (FMRIB’s Software Library, www.fmrib.ox.ac.uk/fsl) using temporal concatenation, to generate independent components shared across all participants, once for pre-assessment and once for post-assessment. We limited the number of components to 20 following previous research (Cushing, Peng, et al., 2023; Webb et al., 2016). This allowed us to limit the number of comparisons and to prevent splitting of components into subcomponents. Group-level functional resting state networks were correlated between pre- and post-assessment using the fslcc function of FSL to identify matching networks between time points. Networks that were correlated above .3 were visually inspected to confirm similarity while thresholded at Z = 3.2, which revealed four candidate matching resting state networks. After group-ICA, we decomposed each group-level component into subject-specific components using dual regression in FSL (Nickerson et al., 2017). Individual participant-specific spatial maps for each of the four networks were subtracted post-treatment to pre-treatment to represent differences between timepoints. To test which networks demonstrated changes in connectivity following neuro-reinforcement, the subtracted networks were subjected to a non-parametric one-sample t-test using FSL randomise with 5000 permutations, threshold-free cluster enhancement, and variance smoothing. Results were corrected for these four comparisons using Bonferroni correction, using a threshold of p < 0.0125. This resulted in one matching network (pre- to post-treatment spatial correlation r = .73) with significant connectivity differences pre- to post-treatment for further analysis.

We created a region of interest out of these significant connectivity changes by taking the corrected p-value map, thresholding at p = .0125, and binarizing. We then extracted the average ICA connectivity estimates from each participant-level spatial map at pre-treatment and post-treatment masked to this region using FSL’s fslmeants, which were then submitted to a linear regression.

### Statistical Analysis

We assessed the relationship between connectivity changes in our resting state network pre-treatment to post-treatment and levels of amygdala activity in response to the target feared animal during the fear test pre-treatment to post-treatment using a linear regression using R (v4.2.2). For analytic consistency with Cushing et al., (2023), we covaried for matching variables with the original analysis. Total number of phobias was included as a covariate to control for variance in the number of diagnosed phobias per person in the sample. Number of neuro-reinforcement sessions was also included as a covariate as we were not sufficiently powered to analyze effects between each dosage group (as a result of COVID-19 shutdowns). Finally, the pre-treatment resting state scans were collected at our initial visit for decoder construction, which was not required to be within a set amount of days before post-treatment. Therefore, we controlled for this variance in time between visits by including time between decoder construction and post-treatment scans in our model.

## RESULTS

The resting state network we analyzed at pre-treatment and post-treatment (Figure 1) encompassed areas broadly implicated in vision and object recognition such as the middle temporal gyrus, lateral occipital cortex, intracalcarine cortex, lingual gyrus, occipital pole, and retrosplenial cortex (Al-Ramadhani et al., 2021; Nagy et al., 2012; Onitsuka et al., 2004; Palejwala et al., 2021; Park-Braswell et al., 2022).

**Figure 1.**
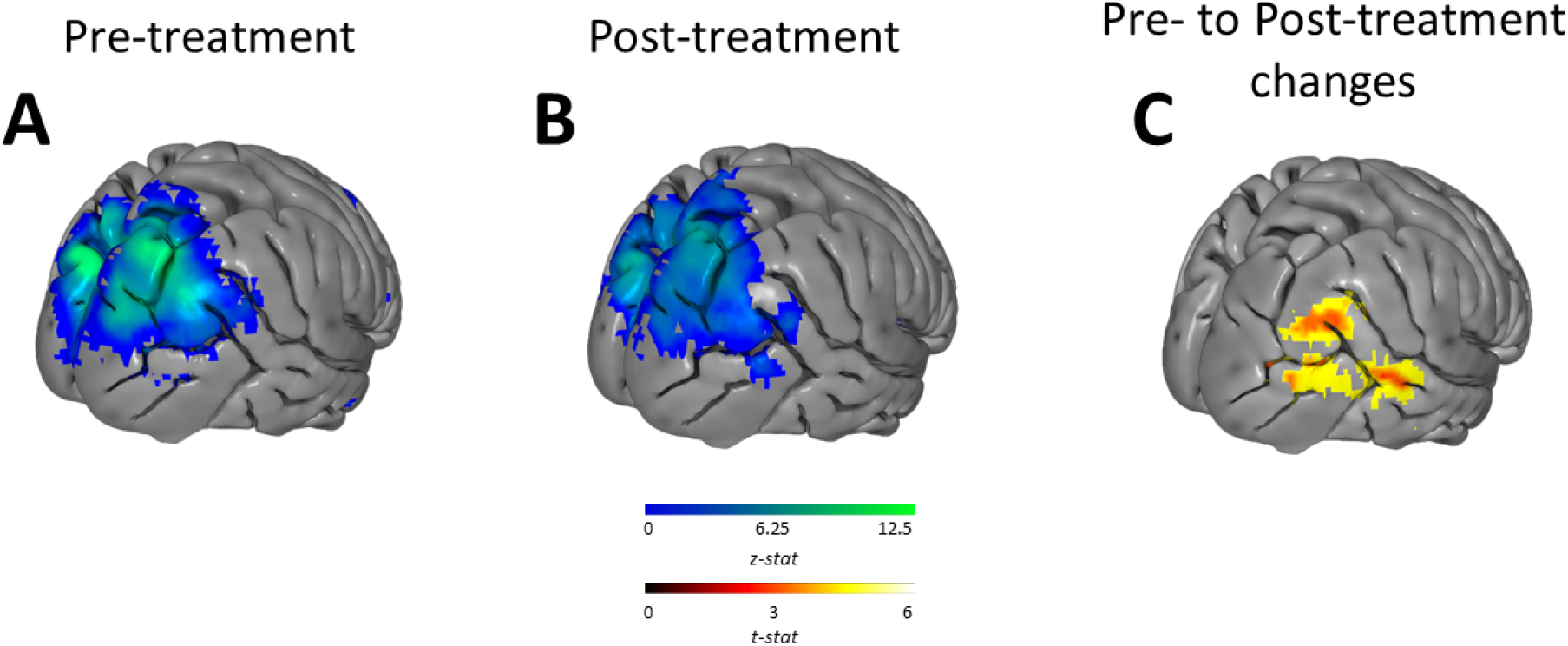
Resting state network before and after multivoxel neuro-reinforcement. A) Map of resting state network activation at pre-treatment (blue/green). B) Map of resting state network activation at post-treatment (blue/green). C) Map of significant increases in resting state network from pre-treatment to post-treatment (red/yellow). Brain images were generated using pyCortex (Gao et al., 2015).

Eight clusters (Table 1) showed significant increase in connectivity pre- to post-treatment. The clusters primarily resided within the middle temporal gyrus, lingual gyrus, supramarginal gyrus, angular gyrus, lateral occipital cortex, retrosplenial cortex, and intracalcarine cortex.

**Table 1.**
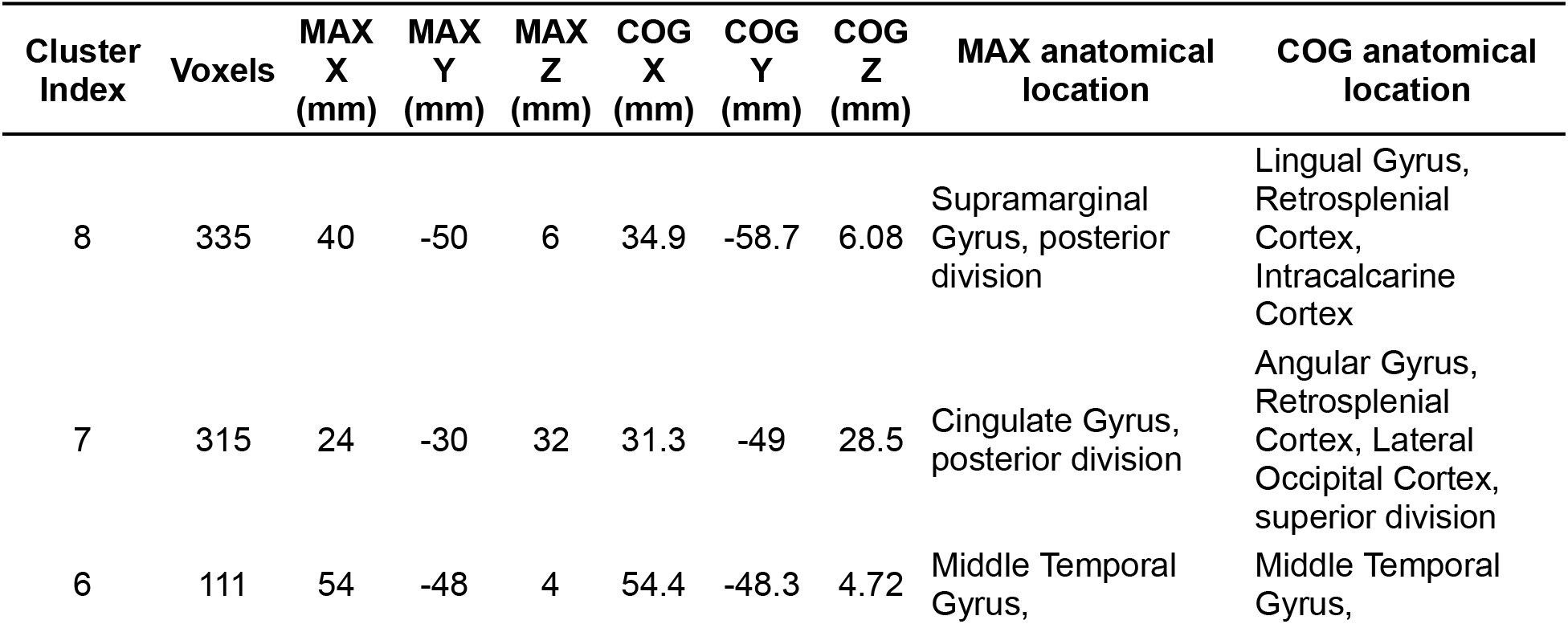
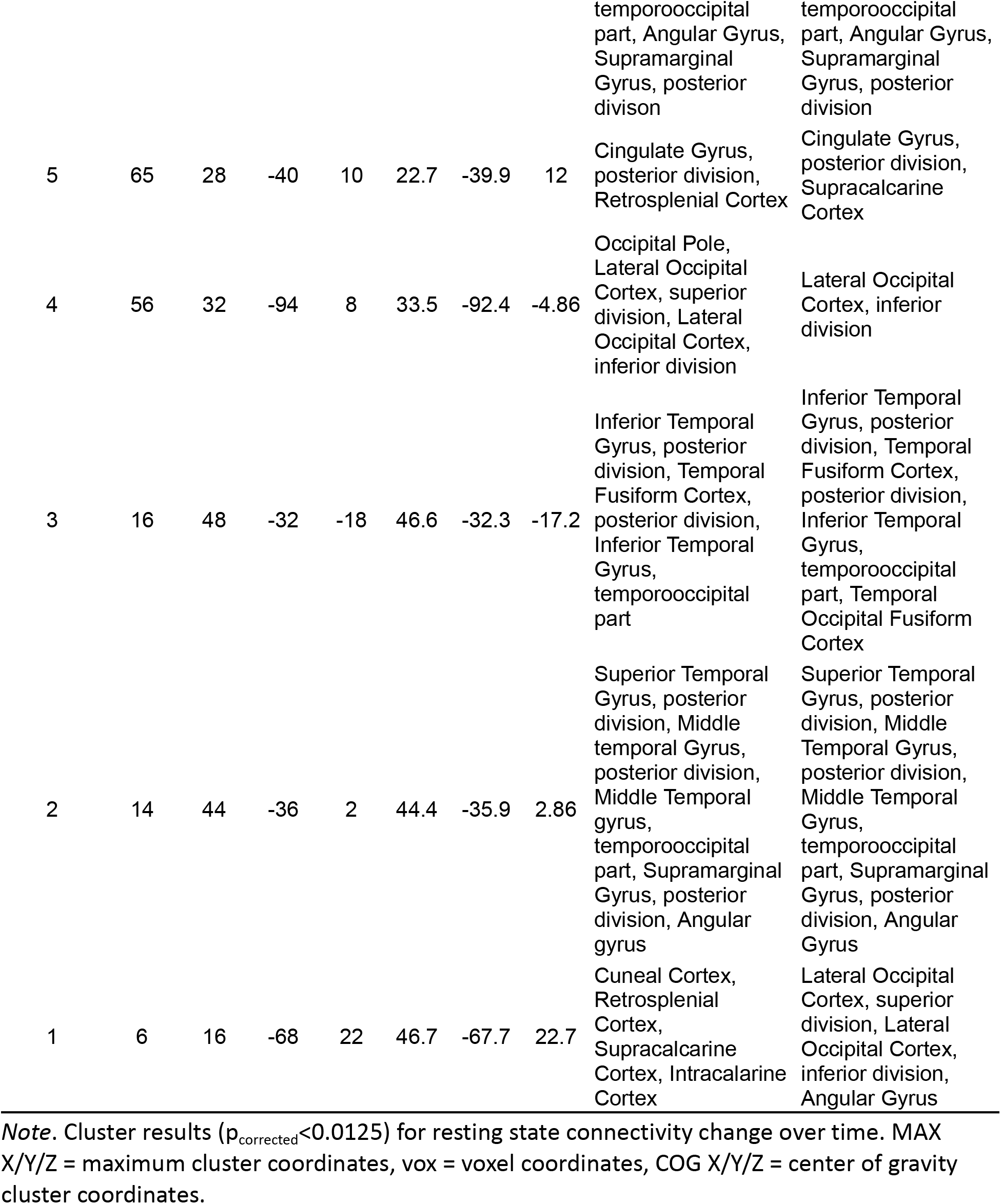
Locations of Clusters of Significant Change of Resting State Connectivity.

| Cluster Index | Voxels | MAX X (mm) | MAX Y (mm) | MAX Z (mm) | COG X (mm) | COG Y (mm) | COG Z (mm) | MAX anatomical location | COG anatomical location |
| --- | --- | --- | --- | --- | --- | --- | --- | --- | --- |
| 8 | 335 | 40 | -50 | 6 | 34.9 | -58.7 | 6.08 | Supramarginal Gyrus, posterior division | Lingual Gyrus, Retrosplenial Cortex, Intracalcarine Cortex |
| 7 | 315 | 24 | -30 | 32 | 31.3 | -49 | 28.5 | Cingulate Gyrus, posterior division | Angular Gyrus, Retrosplenial Cortex, Lateral Occipital Cortex, superior division |
| 6 | 111 | 54 | -48 | 4 | 54.4 | -48.3 | 4.72 | Middle Temporal Gyrus, temporooccipital part, Angular Gyrus, Supramarginal Gyrus, posterior division | Middle Temporal Gyrus, temporooccipital part, Angular Gyrus, Supramarginal Gyrus, posterior division |
| 5 | 65 | 28 | -40 | 10 | 22.7 | -39.9 | 12 | Cingulate Gyrus, posterior division, Retrosplenial Cortex | Cingulate Gyrus, posterior division, Supracalcarine Cortex |
| 4 | 56 | 32 | -94 | 8 | 33.5 | -92.4 | -4.86 | Occipital Pole, Lateral Occipital Cortex, superior division, Lateral Occipital Cortex, inferior division | Lateral Occipital Cortex, inferior division |
| 3 | 16 | 48 | -32 | -18 | 46.6 | -32.3 | -17.2 | Inferior Temporal Gyrus, posterior division, Temporal Fusiform Cortex, posterior division, Inferior Temporal Gyrus, temporooccipital part | Inferior Temporal Gyrus, posterior division, Temporal Fusiform Cortex, posterior division, Inferior Temporal Gyrus, temporooccipital part, Temporal Occipital Fusiform Cortex |
| 2 | 14 | 44 | -36 | 2 | 44.4 | -35.9 | 2.86 | Superior Temporal Gyrus, posterior division, Middle temporal Gyrus, posterior division, Middle Temporal gyrus, temporooccipital part, Supramarginal Gyrus, posterior division, Angular gyrus | Superior Temporal Gyrus, posterior division, Middle Temporal Gyrus, posterior division, Middle Temporal Gyrus, temporooccipital part, Supramarginal Gyrus, posterior division, Angular Gyrus |
| 1 | 6 | 16 | -68 | 22 | 46.7 | -67.7 | 22.7 | Cuneal Cortex, Retrosplenial Cortex, Supracalcarine Cortex, Intracalarine Cortex | Lateral Occipital Cortex, superior division, Lateral Occipital Cortex, inferior division, Angular Gyrus |

*Note.* Cluster results ( $p_{\text{corrected}} < 0.0125$ ) for resting state connectivity change over time. MAX X/Y/Z = maximum cluster coordinates, vox = voxel coordinates, COG X/Y/Z = center of gravity cluster coordinates.

Our reported study (Cushing, Lau, et al., 2023) found significantly decreased amygdala responses for the target animal but not for the control animal during the fear test from pre- to post-treatment. For the current analysis, linear regression was used to investigate the relationship between changes in resting state connectivity and the aforementioned changes in amygdala in response to the target feared animal pre- to post-treatment (Figure 2). Increases in resting state network functional connectivity pre- to post-treatment significantly predicted changes in amygdala pre- to post-treatment for the target animal (**β** = −0.24, t(5) = −3.12, R^2^ = .49, p = .021) but not the control (**β** = −0.24, t(5) = −0.29, R^2^ = .01, p = .782). Greater increases in network connectivity estimates pre-treatment to post-treatment were significantly related to greater decreases in amygdala activation in response to phobic animal stimuli pre-treatment to post-treatment. This aligns with our hypothesis that increases in resting state networks would be associated with decreases in amygdala activity as a direct result of neurofeedback.

**Figure 2.**
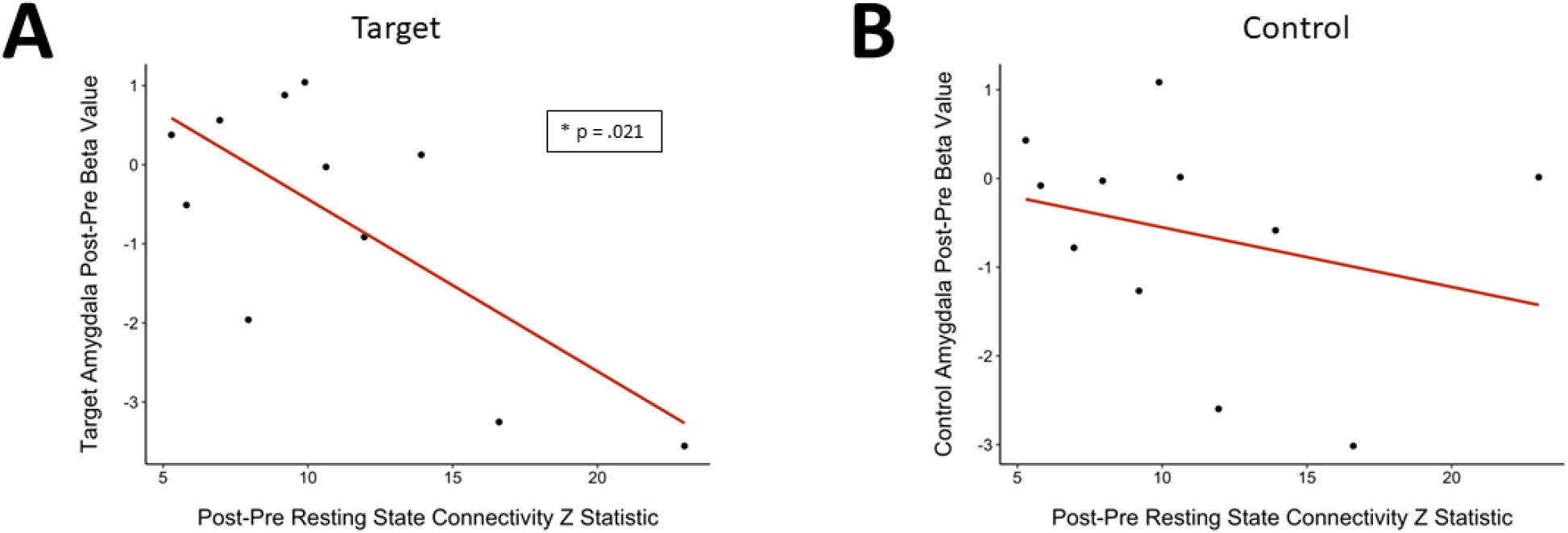
Resting state connectivity changes predict decreased amygdala responding to target phobic stimuli. A) Change in target amygdala response as a function of change in resting state network connectivity pre-treatment to post-treatment. Greater increases in network connectivity post-treatment minus pre-treatment were significantly correlated with greater reductions in amygdala responses post-treatment minus pre-treatment(p < .05). B) Change in control amygdala response as function of change in resting state network connectivity pre-treatment to post-treatment.

## DISCUSSION

In this pilot study, we evaluated changes in resting state functional connectivity following multi-voxel neuro-reinforcement and how they relate to selectively reduced amygdala responses to target phobias following the intervention. We found increases in resting state functional connectivity in regions associated with higher-level visual processing. Increases in resting state functional connectivity were associated with task-based changes in amygdala response to the phobic stimulus that was the target of neuro-reinforcement. Together, these findings from our pilot study preliminarily suggest that these regions could play an intermediary role between reinforcement of animal category-specific multi-voxel patterns in the VT and decreased amygdala activity to feared animals in specific phobia.

First, we identified a functional connectivity network at pre-treatment and post-treatment during rest overlapping with the visual system primarily in the temporal and occipital lobes. This network included areas such as the middle temporal gyrus, lateral occipital cortex, intracalcarine cortex, lingual gyrus, occipital pole, and retrosplenial cortex which are associated with vision and object recognition (Al-Ramadhani et al., 2021; Nagy et al., 2012; Onitsuka et al., 2004; Palejwala et al., 2021; Park-Braswell et al., 2022). Next, we observed significant increases in functional connectivity in the network identified pre- to post-treatment in the parietal, occipital, and temporal lobes. Similarly, these areas are implicated in the categorization of visual information and other higher-order visual processes (Cannon et al., 2007), visio-spatial imagery and processing (Al-Ramadhani et al., 2021; Nakamura et al., 2020), and as well as emotion processing (Stevens et al., 2011). These findings align with a previous information transmission analysis that observed significant transmission during multi-voxel neuro-reinforcement between the VT and the lingual and temporal gyri as well as the fusiform cortex (Taschereau-Dumouchel et al., 2018). Our findings suggest that information transmission during multi-voxel neuro-reinforcement may contribute to changes in functional connectivity during rest. They also demonstrate how repeated reinforcement of multi-voxel patterns in the VT contributes to high-level visual system connectivity outside of the VT during rest. As such, we report novel findings in demonstrating functional connectivity changes in the larger visual system despite targeting pattern activations specifically in the VT for multi-voxel neuro-reinforcement.

Additionally, we observed that increases in functional connectivity were significantly associated with selective decreases in amygdala reactivity to images of the target phobic stimuli in a task. Specifically, the greater the increase in resting state functional connectivity in the visual system pre- to post-treatment, the greater the decrease in task-based amygdala reactivity in response to the target phobic animal pre- to post-treatment. No association was found between increases in functional connectivity and changes in amygdala reactivity to the control animal pre- to post-treatment. Importantly, changes in functional connectivity were isolated away from the amygdala, highlighting that VT multi-voxel neuro-reinforcement does not act on the amygdala directly. This pattern of findings could suggest an inhibitory influence on the amygdala from higher level visual processing regions of the brain through a pathway parallel to an excitatory fear signaling pathway, similar to previous conjectures (Silverstein & Ingvar, 2015). Alternatively, threat detection from the amygdala based on information in the visual system could have decreased through a habituation effect, where repeated activations of multi-voxel animal patterns in the VT led to a diminished threat detection response from the amygdala when viewing feared stimuli.

A primary strength in this analysis is the sample of participants with diagnosed animal phobias, as this is the first study of multi-voxel neuro-reinforcement to investigate changes in a population of this severity of animal phobia. These findings further validate the possibility of multi-voxel neuro-reinforcement as a clinical intervention by highlighting the possibility for persistent changes in brain function that could be responsible for its efficacy. Because our group-ICA approach was model-free while utilizing non-parametric statistics, we were able to detect significant results at a whole-brain level using robust methods despite our limited sample size. Additionally, observing increases in functional connectivity in regions related to the visual system in the absence of a predetermined model further supports the robustness of our data-driven findings.

However, the study is not without limitations. First, our sample size was relatively small due to inherent difficulties with recruitment stemming from the COVID-19 pandemic as well as technical issues and scanning time limitations that led to incomplete scans as the resting state scans were a secondary measure in the study. Second, the study did not include a between-subjects control group. However, we applied control conditions within subjects, which ultimately allowed us to achieve a double-blinded design at higher statistical power than a between-subject design. Future studies should examine these effects in the context of a between-subjects design.

Last, the meaning of these observed changes in functional connectivity is still underdetermined. In the future, more targeted investigations of the brain regions that demonstrated changes can help identify the direct connections that these high level visual processing regions have with both the VT and the amygdala. Considering the pilot nature of our investigation, future studies should also seek to replicate these analyses with a larger clinical sample to increase statistical power and reveal more potential effects that may have been hidden in our sample. These results provide a set of brain regions that are demonstrated to change as a result of multi-voxel neuro-reinforcement for future studies to further investigate in order to delineate specific causal pathways between high level areas of the visual system, the VT, and the amygdala.

Overall, the study found significant increases in functional connectivity in the visual system as well as a significant relationship between increased functional connectivity strength and decreases in amygdala reactivity in response to the target animal. These changes could suggest high level visual processing areas as a potential bridge that explains the connection between the VT area and the amygdala. These findings from the first randomized clinical trial of multi-voxel neuro-feedback for animal phobia also represent the early steps in elucidating mechanisms of change for this yet understudied topic. With further research building atop these results and a greater understanding of its mechanisms, this novel treatment may open up fruitful new approaches to treating phobia and anxiety disorders.

## Data Availability

All data produced in the present study are available upon reasonable request to the authors.

## Acknowledgments

The authors would like to thank study coordinators Ana Costello and Annelise Murillo for their assistance to this study through participant recruitment and data collection.

HL received financial support from the US National Institute of Mental Health (R61MH113772) and The Templeton World Charity Foundation (RA537-01). MK was supported by AMED under grant number JP18dm0307008. MGC received financial support from the US National Institute of Mental Health (R61MH113772). VT-D received financial support from the Fonds de Recherche du Québec - Santé (FRQS).

